# LAMP-BEAC: Detection of SARS-CoV-2 RNA Using RT-LAMP and Molecular Beacons

**DOI:** 10.1101/2020.08.13.20173757

**Authors:** Scott Sherrill-Mix, Young Hwang, Aoife M. Roche, Abigail Glascock, Susan R. Weiss, Yize Li, Leila Haddad, Peter Deraska, Caitlin Monahan, Andrew Kromer, Jevon Graham-Wooten, Louis J. Taylor, Benjamin S. Abella, Arupa Ganguly, Ronald G. Collman, Gregory D. Van Duyne, Frederic D. Bushman

## Abstract

**Background:** Rapid spread of SARS-CoV-2 has led to a global pandemic, resulting in the need for rapid assays to allow diagnosis and prevention of transmission. Reverse Transcription-Polymerase Chain Reaction (RT-PCR) provides a gold standard assay for SARS-CoV-2 RNA, but tests are expensive and supply chains are potentially fragile, motivating interest in additional assay methods. Reverse Transcription and Loop-Mediated Isothermal Amplification (RT-LAMP) provides an alternative that uses orthogonal and often less expensive reagents without the need for thermocyclers. The presence of SARS-CoV-2 RNA is typically detected using dyes to report bulk amplification of DNA; however, a common artifact is nonspecific DNA amplification, which complicates detection.

**Results:** Here we describe the design and testing of molecular beacons, which allow sequence-specific detection of SARS-CoV-2 genomes with improved discrimination in simple reaction mixtures. To optimize beacons for RT-LAMP, multiple locked nucleic acid monomers were incorporated to elevate melting temperatures. We also show how beacons with different fluorescent labels can allow convenient multiplex detection of several amplicons in “single pot” reactions, including incorporation of a human RNA LAMP-BEAC assay to confirm sample integrity. Comparison of LAMP-BEAC and RT-qPCR on clinical saliva samples showed good concordance between assays. To facilitate implementation, we developed custom polymerases for LAMP-BEAC and inexpensive purification procedures, which also facilitates increasing sensitivity by increasing reaction volumes.

**Conclusions:** LAMP-BEAC thus provides an affordable and simple SARS-CoV-2 RNA assay suitable for population screening; implementation of the assay has allowed robust screening of thousands of saliva samples per week.

## Background

Since its first detection in March 2020, the beta-coronavirus SARS-CoV-2 has spread around the world, infecting over 100 million people and causing over 2 million deaths. Frequent asymptomatic spread of this virus means that frequent, rapid and affordable screening and surveillance testing is essential to controlling this pandemic [1-3].

Numerous methods have been developed to detect SARS-CoV-2 infection. The most common method is RT-qPCR to detect SARS-CoV-2 RNA[4]. RT-qPCR has the advantage of providing accurate and sensitive detection, but supply chain issues have at times limited testing, motivating the development of additional methods using orthogonal materials. RT-LAMP has been widely studied as an alternative[5-9]. LAMP assays use a “rolling hairpin” mechanism to allow amplification at a single temperature using polymerase enzymes different from those used for PCR, helping avoid supply chain bottle necks. In addition, RT-LAMP can be implemented on neat saliva, or on RNA purified using simple reagents available in bulk[9], again helping bypass supply chain issues and adding robustness to assays.

RT-LAMP assays are typically not as sensitive as RT-qPCR assays[1], but the importance of this varies with the application. Clinical diagnostic tests typically require high sensitivity; however, studies suggest that infected individuals are far more infectious during periods of peak viral loads, so methods for screening asymptomatic populations can be adequate even with lesser sensitivities [1, 2]. A recent study emphasized that frequency of testing and speed of reporting results are much more important than assay sensitivity for reducing transmission, emphasizing the value of assays like RT-LAMP that may be implemented efficiently and inexpensively[1].

However, a complication is that RT-LAMP reactions often result in non-specific amplification in the absence of target, particularly at longer reaction times, limiting sensitivity. This off-target amplification is especially problematic because LAMP reactions are commonly quantified using colorimetric or fluorescent dyes reporting only bulk DNA synthesis. To address these problems, improvements based on sequence-specific detection have been proposed such as incorporating DNA sequencing (LAMP-seq)[10] or CAS enzymes (DETECTR)[11]. These methods are promising, but as presently designed they typically require opening of RT-LAMP tubes and secondary manipulation of reaction products, which has the potential to result in contamination of subsequent reactions with amplification products from previous assays. In another detection method, a quencher-fluorophore duplex can be created by adding a short oligonucleotide complementary to a standard LAMP primer which is then displaced upon amplification[12-15]. These methods are more specific than bulk reporters but are still potentially vulnerable to false positives from spurious amplification.

Previous research has shown the potential for molecular beacons[16] to allow sequence-specific detection of LAMP products in “single-pot” assays [17, 18]. Here, we adapt molecular beacons to detect SARS-CoV-2 sequences, a method we have named LAMP-BEAC (Fig. 1a). Molecular beacons are target-specific oligonucleotides labeled with a fluorophore on one end and a quencher on the other. The beacons are designed to incorporate complementary sequences on their 5’ and 3’ ends such that at low temperatures the ends anneal to form a hairpin, bringing the quencher and fluorophore into close proximity and quenching fluorescence. When the target of interest is present, the complementary target-specific beacon sequence anneals to its target, separating the fluorophore from the quencher and greatly increasing the fluorescent signal. The binding sites for beacons can be targeted to amplicon sequences not present in oligonucleotides used for priming, thereby enhancing specificity. The increase in fluorescence resulting from annealing of the beacon probe can be detected without manipulation of the product or opening the reaction tube. Here we describe 1) development of molecular beacons for detection of SARS-CoV-2 RNA in LAMP-BEAC reactions, 2) development of a LAMP-BEAC method to detect human RNA to validate sample integrity, 3) combinations of LAMP-BEAC assays for single-pot multiplex detection, 4) development of custom polymerases allowing inexpensive expression and purification of required enzymes, 5) use of LAMP-BEAC to screen infected subjects for viral RNA in saliva, and 6) increased sensitivity accessible using the high specificity of molecular beacons.

**Fig. 1.**
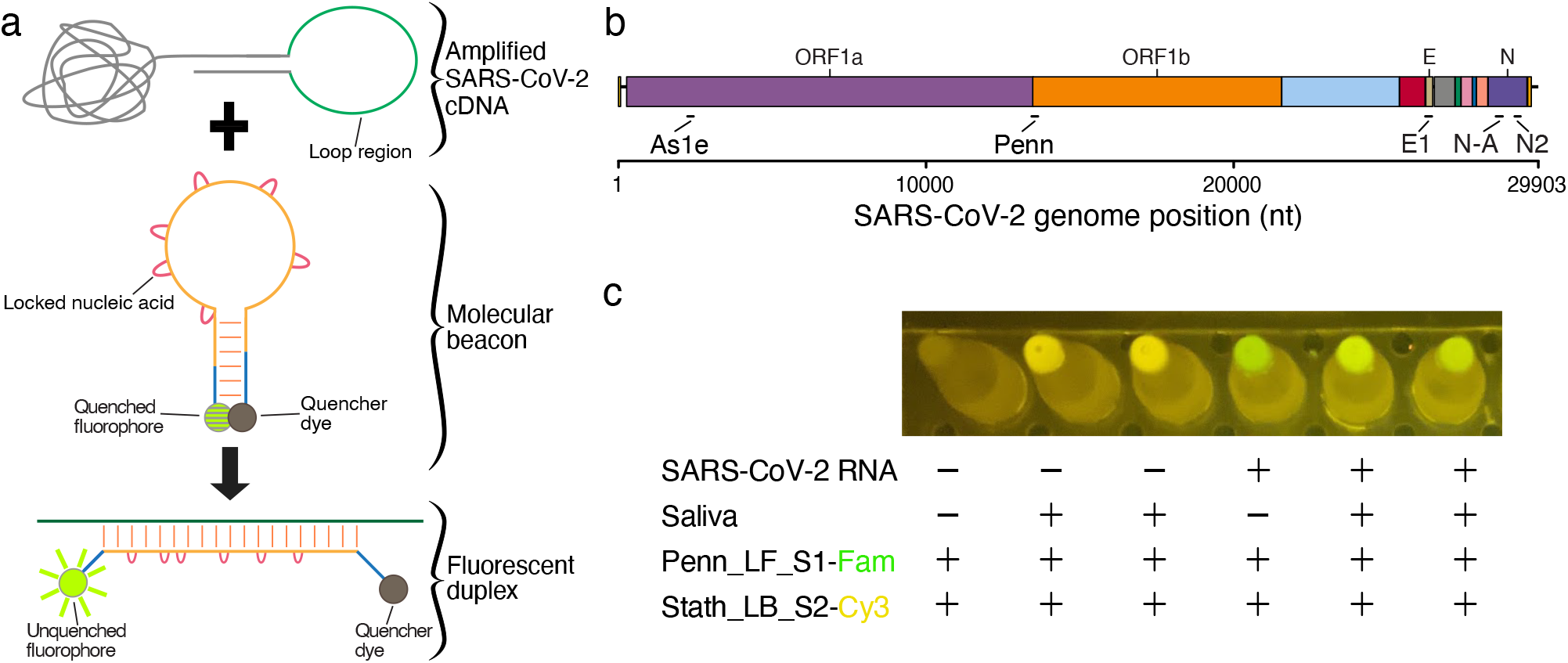
LAMP-BEAC: RT-LAMP assayed using molecular beacons. A) Product from a LAMP reaction is depicted emphasizing its loop region which forms single stranded loops during amplification along with an example molecular beacon in its annealed hairpin form, which is quenched. Binding of the beacon to the target complementary sequence LAMP amplicon separates the fluorescent group and the quencher, allowing detection of fluorescence. The red loops on the beacon indicate locked nucleic acids used to increase binding affinity. B) A genome map of SARS-CoV-2 showing the locations of primer binding sites for the LAMP primer sets used in this study. C) Example of visual detection of LAMP-BEAC fluorescence using an orange filter with blue illumination. Multiplexed SARS-CoV-2 targeted Penn and human control STATH primer sets were used to amplify samples consisting of water or inactivated saliva with or without synthetic SARS-CoV-2 RNA (10,000 copies per reaction). Molecular beacons Penn_LF_S1 conjugated to a FAM fluorophore fluorescing green and Stath_LB_S2 conjugated to Cy3 fluorescing yellow were included in the reaction. The image was captured using the “Night Sight” mode of a Google Pixel 2 cell phone.

## Results

### Designing molecular beacons for SARS-CoV-2 RT-LAMP

Several beacons were tested for detection of SARS-CoV-2 RNA in RT-LAMP reactions (Table S1). Optimization required identifying sequence designs that performed properly under the conditions of the RT-LAMP reaction, which is typically run at temperatures around 65°C. Function of the beacon requires that the hairpin remain mostly folded in the hairpin structure at this temperature, while still opening sufficiently often to allow annealing to the target RT-LAMP cDNA product. The annealed beacon-target cDNA duplex must then be sufficiently stable at 65°C to result in unquenching and an increase in fluorescence. To increase beacon affinity for use at higher temperatures, we substituted multiple dNTP positions within the target sequence of each beacon with locked nucleic acids [17]. Locked nucleic acids reduce the conformational flexibility of dNTPs and make the free energy of nucleic acid annealing more favorable [19]. We tested the performance of 28 molecular beacons using five previously reported SARS-CoV-2 (Fig. 1b) and three human control RT-LAMP amplicons (Fig. S3, Table S1).

### Testing LAMP-BEAC

An example of a successful beacon design is Penn_LFMB_S1 (Table S1). The RT-LAMP amplicon targets the orf1ab coding region and was first reported by El-Tholoth and coworkers at the University of Pennsylvania (named “Penn”)[7]. The favored beacon was designed to target sequences within the forward DNA loop generated during LAMP; thus the beacon is designated Penn loop forward beacon, contracted to Penn_LFMB_S1. Detection can be accomplished with laboratory plate readers or PCR machines (below), and even visually with a simple blue light and orange filter (Fig. 1c).

Figure 2 shows use of the Penn_LFMB_S1 system to detect synthetic SARS-CoV-2 RNA. Tests were carried out with commercial LAMP polymerase and reverse transcriptase preparations. In addition, to avoid possible supply chain problems and allow potential production of reagents in resource limited settings, we produced and purified novel DNA polymerase and reverse transcriptase enzymes, which were assayed in parallel with commercial preparations for some tests (described below).

**Fig. 2.**
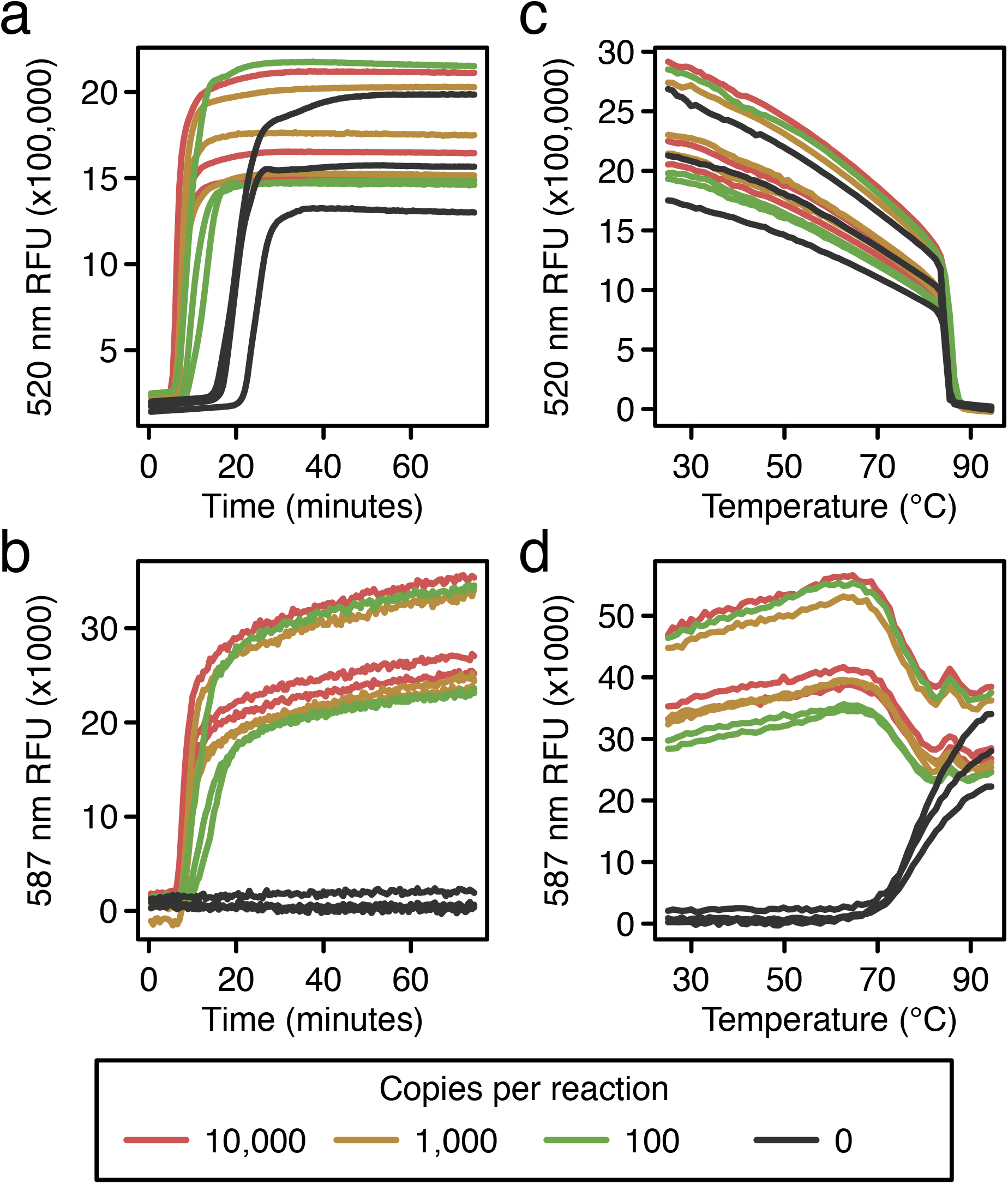
Reaction progression curves comparing RT-LAMP using the Penn primer set assayed using an intercalating dye and the Penn_LFMB_S1 molecular beacon in the same reactions. A) Conventional RT-LAMP assay using non-specific dye to detect amplification of synthetic SARS-CoV-2 RNA diluted in water. Time after reaction initiation (x-axis) is compared to the relative fluorescence intensity (y-axis). The copy numbers of SARS-CoV-2 RNA in the reaction mixtures are shown in the key at the bottom. B) Detection of the amplification of SARS-CoV-2 RNA using a LAMP-BEAC molecular beacon in the same reactions shown in A. Lines are colored as in A. C) and D) Thermal melting curves to characterize amplification products. The results shown are for the same reactions as in A and B. Reaction products were cooled to room temperature, then slowly heated for the melt curve analysis. C) Characterization of the fluorescence intensity produced by non-specific intercalating dye (y-axis) with RT-LAMP end products over varying temperatures (x-axis). Lines are colored as in A. D) Characterization of the fluorescence intensity produced by a LAMP-BEAC molecular beacon with RT-LAMP end products over varying temperatures. Markings as in C.

To compare standard LAMP amplification with LAMP-BEAC, reactions were prepared containing both fluorescent dye (Fig. 2a), which detects bulk DNA by intercalation, and the molecular beacon Penn_LFMB_S1 (Fig. 2b). Reaction products were detected at two wavelengths, allowing separate quantification of the bulk dye and the molecular beacon in single reactions. The non-specific intercalating dye reported bulk DNA production in positive samples earlier than the water controls, but the negative controls did amplify shortly after. This spurious late amplification is commonly seen with RT-LAMP, though the mechanism is unclear. The primers may interact with each other to form products and launch amplification, or perhaps the reaction results from amplification of adventitious environmental DNA. In separate tests, synthesis of DNA products was shown to depend on addition of LAMP primers (data not shown).

Molecular beacon Penn_LFMB_S1 in the same reactions showed more clear-cut discrimination (Fig. 2b). The positive samples showed positive signal, but no signal was detected for the negative water controls. Lack of amplification in negative controls has been reproducible over multiple independent reactions (examples below).

The nature of the products could be assessed using thermal denaturation (Fig. 2c and d). Reactions were first cooled to allow full annealing of complementary DNA strands, then slowly heated while recording fluorescence intensity. The fluorescent signal of the intercalating dye started high but dropped with increasing temperature in all samples (Fig. 2c), consistent with denaturation of the duplex and release of the intercalating dye into solution. In contrast, the beacon’s fluorescent signal in the water controls started at low fluorescence (Fig. 2d), consistent with annealing of the beacon DNA termini to form the hairpin structure (Fig. 1a). At temperatures above 70 °C, the fluorescence modestly increased, consistent with opening of the hairpin and reptation of the beacon as a random coil in solution. For reactions containing the RT-LAMP product and Penn_LFMB_S1 beacon, fluorescence values were high at lower temperatures, consistent with formation of the annealed duplex, then at temperature sufficient for denaturation, the fluorescence values fell to match those of the random coil (Fig. 2d). Thus, the LAMP-BEAC assay generates strong fluorescence signals during LAMP amplification in the presence of target RNA but not in negative controls, and the thermal melting properties are consistent with formation of the expected products.

### Multiplex LAMP-BEAC assays

We next sought to develop additional LAMP-BEAC assays to allow multiplex detection of SARS-CoV-2 RNA, and to allow parallel analysis of human RNA controls as a check on sample integrity, and so developed several additional beacons (Supplementary Table 1). E1_LBMB_S1 recognizes an amplicon targeting the viral E gene reported in [20], As1e_LBMB_S2 recognizes the As1e amplicon reported in [9] targeting the orf1ab coding region, N2_LBMB_S3 recognizes N2 amplicon reported in [20] targeting the N coding region, and N-A_LFMB_S2 recognizes N-A amplicon reported in [21] targeting the N coding region (Fig. 1b). We also developed positive control beacons, STATH_LFMB_S1 and a later brighter iteration STATH_LBMB_S2, to detect a LAMP amplicon targeting the human statherin mRNA [22]. Additional control beacons included ACTB to detect beta-actin mRNA [20] and RNaseP to detect ribonuclease P subunit p20 POP7 mRNA or DNA [23] (Supplementary Table 1, Fig. S3). We chose to focus on STATH for further testing because it is abundantly expressed in human saliva and spans an exon junction to allow selective detection of RNA and not DNA.

To allow independent detection of each amplicon as a quadruplex assay, each beacon was labeled using fluorophores with different wavelengths of maximum emission. For example, E1_LBMB_S1 was labeled with FAM and detected at 520 nm, STATH_LFMB_S1 was labeled with hexachlorofluorescein (Hex) and detected at 587 nm, As1e_LBMB_S2 was labeled with Tex615 and detected at 623 nm, and Penn_LFMB_S1 was labeled with cyanine-5 (Cy5) and detected at 682 nm.

This quadruplex LAMP-BEAC assay was tested with contrived samples, in which saliva was doped with synthetic SARS-CoV-2 RNA (Fig. 3). Prior to dilution, saliva was treated with TCEP and EDTA, followed by heating at 95 °C, which inactivates both SARS-CoV-2 and cellular RNases[9], and so is part of our sample processing pipeline. The STATH_LFMB_S1 amplicon detected the human RNA control in all saliva samples (Fig. 3a).The E1_LBMB_S1 and As1e_LBMB_S2 amplicons both consistently detected SARS-CoV-2 RNA down to ∼250 copies per reaction(Fig. 3bc). Samples were called positive if either E1 or As1e showed amplification. Using this scoring method, the combination consistently detected SARS-CoV-2 down to 125 copies, and even detected 2/3 positives at 16 copies per reaction.

**Fig. 3.**
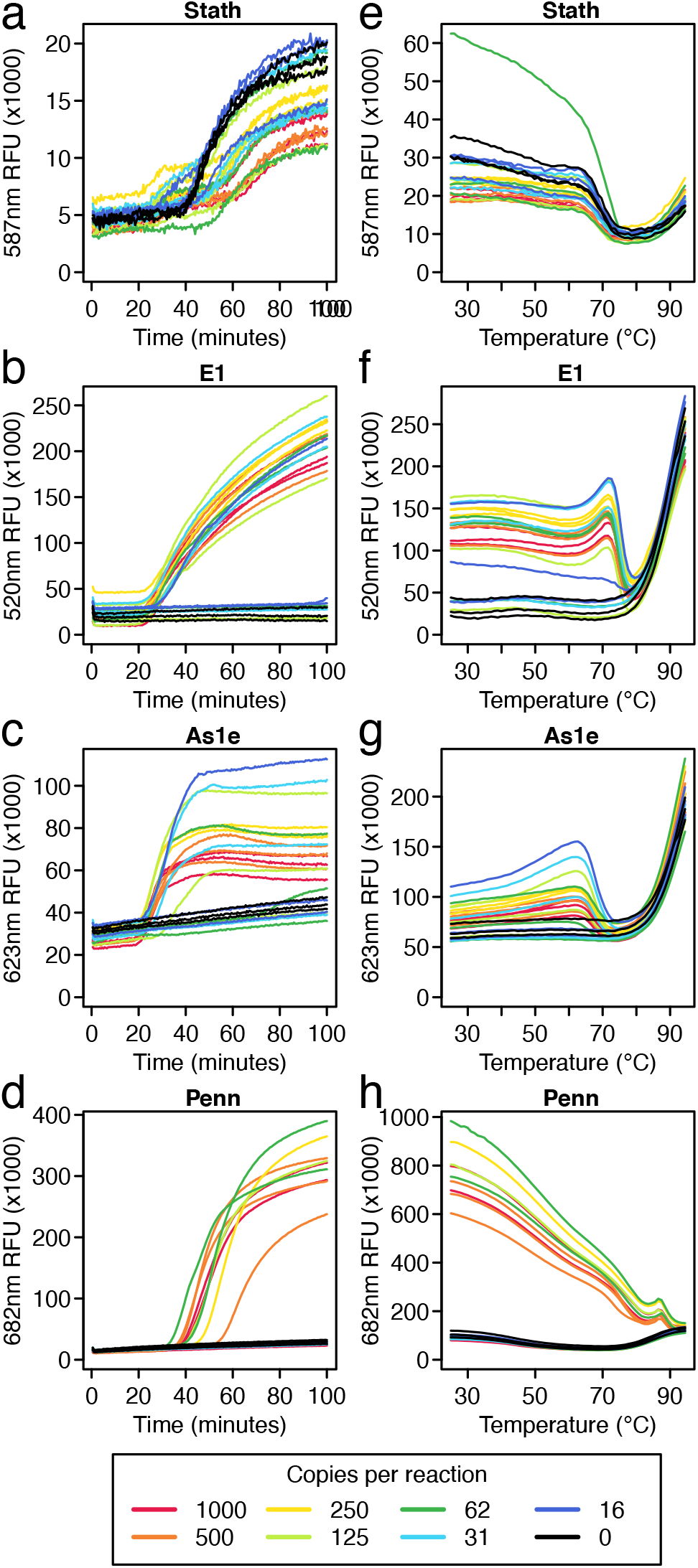
A multiplex LAMP-BEAC method assaying four amplicons. Assays were carried out using a LAMP primers and molecular beacon to detect human STATH RNA (A) and three primer sets and corresponding beacons to detect SARS-CoV-2: E1 (B), As1e (C), and Penn (D). For these assays, synthetic SARS-CoV-2 RNA was diluted into saliva (inactivated as described[9]); copies per reaction are shown by the color code below the figures. A-D) Amplification curves of the reactions showing fluorescent intensity (x-axis) over time (y-axis). E-H) Endpoint melt curves of the reactions shown in A-D showing changes in fluorescent intensity (y-axis) as the temperatures (x-axis) of the final reaction products were raised from 25 to 95 °C.

The Penn_LFMB_S1 amplicon was least sensitive, detecting SARS-CoV-2 RNA consistently only at ∼1000 copies per reaction (Fig. 3d). In the multiplex setting, the Penn amplicon sensitivity was lower than that observed when run in isolation (Fig. 2), likely indicating competition between amplicons during multiplexed reactions. Thus use of the Penn_LFMB_S1 assay in the multiplex format selectively reports particularly high RNA copy numbers.

A useful feature of the STATH control amplicon used here is that it amplifies more slowly than the SARS-CoV-2 amplicons. Slower amplification of human controls is desirable to avoid exhaustion of reaction components due to competition, which could prevent viral detection.

Melt curve analysis was also carried out to verify reaction products (Fig. 3 e-h). Melt curve profiles were distinctive for each beacon, but the overall pattern included high fluorescence in the positive samples and low values in negative samples at lower temperatures, then convergence of positive and negative samples at high temperatures associated with full melting of the beacon and reptation in solution. The melt curve data for each beacon supported correct function and the expected structures of the amplification products.

### Assessing LAMP-BEAC performance on clinical saliva samples

We next tested the LAMP-BEAC assay on a set of 82 saliva samples collected during surveillance for potential SARS-CoV-2 infection. Samples were from a clinical site, where subjects were tested by nasopharyngeal (NP) swabbing and clinical RT-qPCR, and also donated saliva for comparison. Saliva samples were treated with TCEP and EDTA and heated at 95 °C for five minutes to inactivate RNase and SARS-CoV-2 [9]. We performed a triplex LAMP-BEAC assay using Penn_LFMB_S1, N2_LBMB_S3 and STATH_LBMB_S2 beacons in a set of five 20 μl reactions. As an additional check, RNA was purified from the same set of saliva samples and RT-qPCR carried out using the CDC-recommend N1 primer set.

Absence of STATH amplification can indicate potential RNA degradation or inhibitors in the sample, but another reason for lost signal can be competition between amplicons. STATH tends to amplify more slowly than the SARS-CoV-2-targeted amplicons, and so can be suppressed by robust amplification of viral amplicons (Figure 3). In practice, we suggest that a sample with SARS-CoV-2 amplification should be called as positive regardless of STATH results, a sample with no SARS-CoV-2 amplification should be called as negative if STATH amplification is observed, and a sample with no STATH amplification and no viral amplification should be called indeterminant.

In these clinical samples, some degradation was apparent and STATH amplification was detected in only 56 out of 82 samples (Supplementary Figure S1, Supplementary Table S2). These saliva samples had been stored for months and frozen and thawed multiple times, so some attrition is not surprising. SARS-CoV-2 amplification was observed in 6 of these STATH failures suggesting potential competition between amplicons or a greater robustness of viral RNA. Where STATH or SARS-CoV-2 amplification was detectable, the LAMP-BEAC assay correlated perfectly with the amplification of SARS-CoV-2 above the limit of detection by laboratory RT-qPCR on the same saliva samples, i.e. a sensitivity and specificity of 1 (Fig. 4). Performance was similar in quadruplex and duplex LAMP-BEAC assays using Penn_LFMB_S1, E1_LBMB_S1, As1e_LBMB_S2 and STATH_LFMB_S1 performed on subsets of the same samples (Supplementary Table S2).

**Fig. 4.**
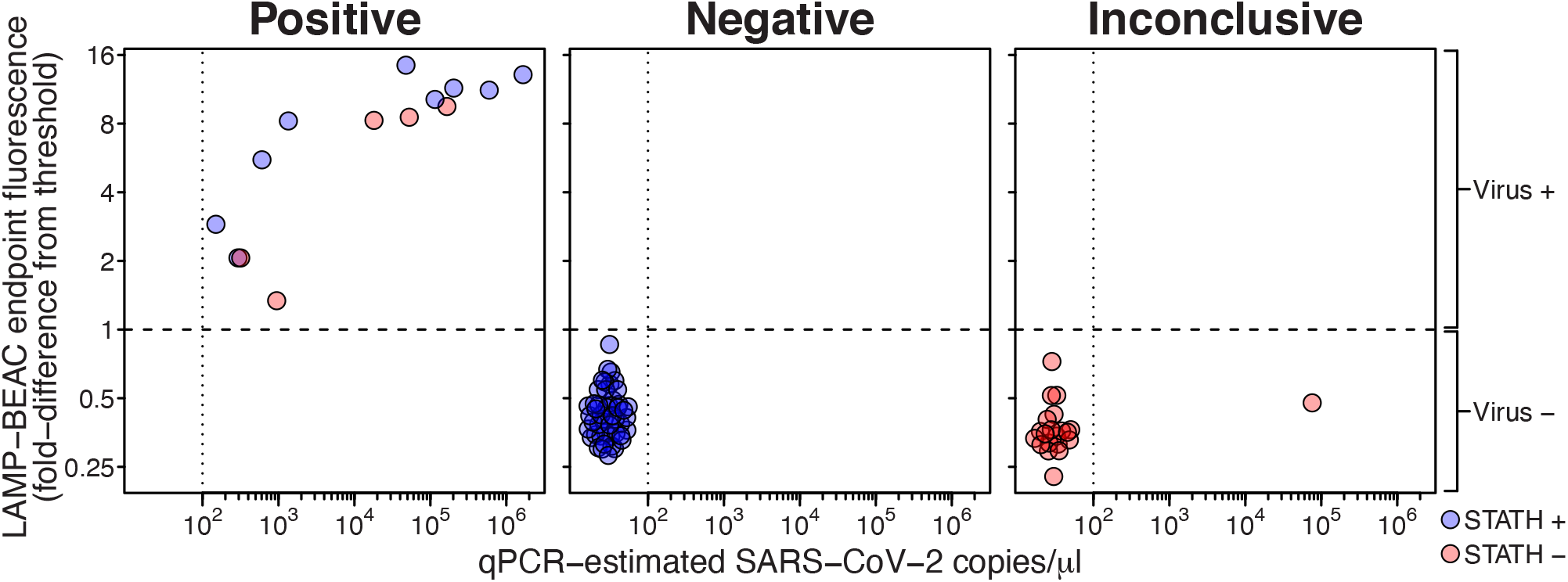
Validation of multiplexed LAMP-BEAC on 82 clinical saliva samples. Inactivated saliva samples were assayed by LAMP-BEAC using molecular beacons STATH_LBMB_S2, N2_LBMB_S3 and Penn_LFMB_S1. Samples were called “Positive” if they had detectable amplification in any SARS-CoV-2 amplicon; if human control STATH amplification was detected but not SARS-CoV-2 they were called “Negative”; if no STATH or SARS-CoV-2 amplification was detected they were called “Inconclusive”. SARS-CoV-2 targeted N2 and Penn fluorescence were quantified as the fold difference from a threshold set at two times the highest fluorescence observed in negative controls (dashed line). The maximum Penn or N2 fluorescence observed among the 5 reactions for each sample (y-axis) was compared to viral copy numbers estimates by laboratory qPCR (x-axis). Samples with qPCR copy numbers below the inferred limit of detection of 100 copies per μl are arbitrarily spaced apart for visualization. Sample integrity was assessed as STATH end point fluorescence greater than 120% the greatest fluorescence observed in water controls. Blue points indicate samples with detected STATH amplification and red indicates samples with no detectable STATH amplification, i.e. potentially indicating degraded samples or competition between amplicons.

Comparison to the results of clinical RT-qPCR testing on NP swabs from the same patients was complicated by disagreements with the laboratory RT-qPCR testing on matched saliva samples. Of the 24 samples scored as positive by clinical testing on NP swabs, only 17 had detectable amplification by laboratory qPCR on saliva and an additional 9 samples with detectable amplification by laboratory qPCR had been marked negative by clinical testing. All but one disagreement (see below) occurred in samples with concentrations inferred as less than 100 copies per μl by laboratory qPCR (clinical quantifications were not available). We thus inferred the laboratory qPCR had a practical limit of detection of 100 copies per μl. The LAMP-BEAC assay did not detect amplification in any of these discrepant samples. For samples with greater than 100 inferred copies per μl, the clinical test results and LAMP-BEAC agreed perfectly with the exception of a single saliva sample called positive by LAMP-BEAC but negative by clinical NP testing. This sample was also estimated at 200,000 viral RNA copies per μl by laboratory RT-qPCR and as positive in 23 LAMP-BEAC amplifications in 14 separate reactions across 4 different primer sets (Supplementary Table S2). A recent study has documented differences between the loads of SARS-CoV-2 RNA at different body sites [24], including oral and nasal sites, potentially accounting at least in part for the observed differences.

We note that the detection shown in Figure 4, using end point fluorescence values and not reaction progression curves, offers a simplified read out for reaction results. That is, advanced qPCR machines are not needed for amplification or quantification of product formation using LAMP-BEAC, but rather reactions can be performed using a simple heat block or incubator and reaction end points can be read out using a simpler fluorescent plate reader or even visual/cell phone detection (Fig. 1c). This may help bypass possible supply chain bottlenecks and expenses associated with purchasing qPCR machines for SARS-CoV-2 assays.

### Laboratory-based production of polymerases required for RT-LAMP

Polymerase enzymes are expensive and potentially subject to supply chain disruptions, so we engineered novel reverse transcriptase and DNA polymerase enzymes and devised simple purification protocols, allowing inexpensive local production of the required enzymes. HIV-2 reverse transcriptase and the polA large fragment from *Geobacillus stearothermophilus* were each engineered to contain several amino acid substitutions expected to stabilize enzyme folding at higher temperatures (RT) or improve strand displacement activity (Bst). Enzymes were purified and tested as described in the methods. Figure S2 summarizes results of side-by-side assays using lab-purified polymerases and commercial enzyme preparations, which indicate that our novel polymerase enzymes are at least as efficient as commercial preparations.

### Increased sensitivity through increased reaction volume

The combination of affordable enzyme and low probability of false positives suggests that it could be possible to increase testing sensitivity by increasing reaction volume and sample input. To test this, we quantified sensitivity versus reaction volume using the N2 primer set, comparing detection with nonspecific dye and the N2_LBMB_S3 beacon.

To test the relationship of reaction volume and sensitivity, we first compared the performance of 10 μl reactions with 4 μl of saliva input versus 20 μl reactions with 8 μl of the same saliva input. Samples were contrived using varying concentrations of synthetic SARS-CoV-2 RNA in inactivated saliva. We observed that the larger 20 μl reaction volume and correspondingly larger saliva input increased detection rate for the molecular beacon and non-specific dye (Fig. 5a-f). However, interpretation of results by non-specific dye was complicated by the variable distribution of cycle thresholds observed in negative controls, making a clean distinction of positive amplification difficult. In contrast, the LAMP-BEAC molecular beacon detected no sequence-specific amplification in any negative sample. The clear binary threshold provided by molecular beacons simplifies interpretation, enables endpoint detection and suggests that sensitivity could be heightened by further increasing reaction volume.

**Fig 5.**
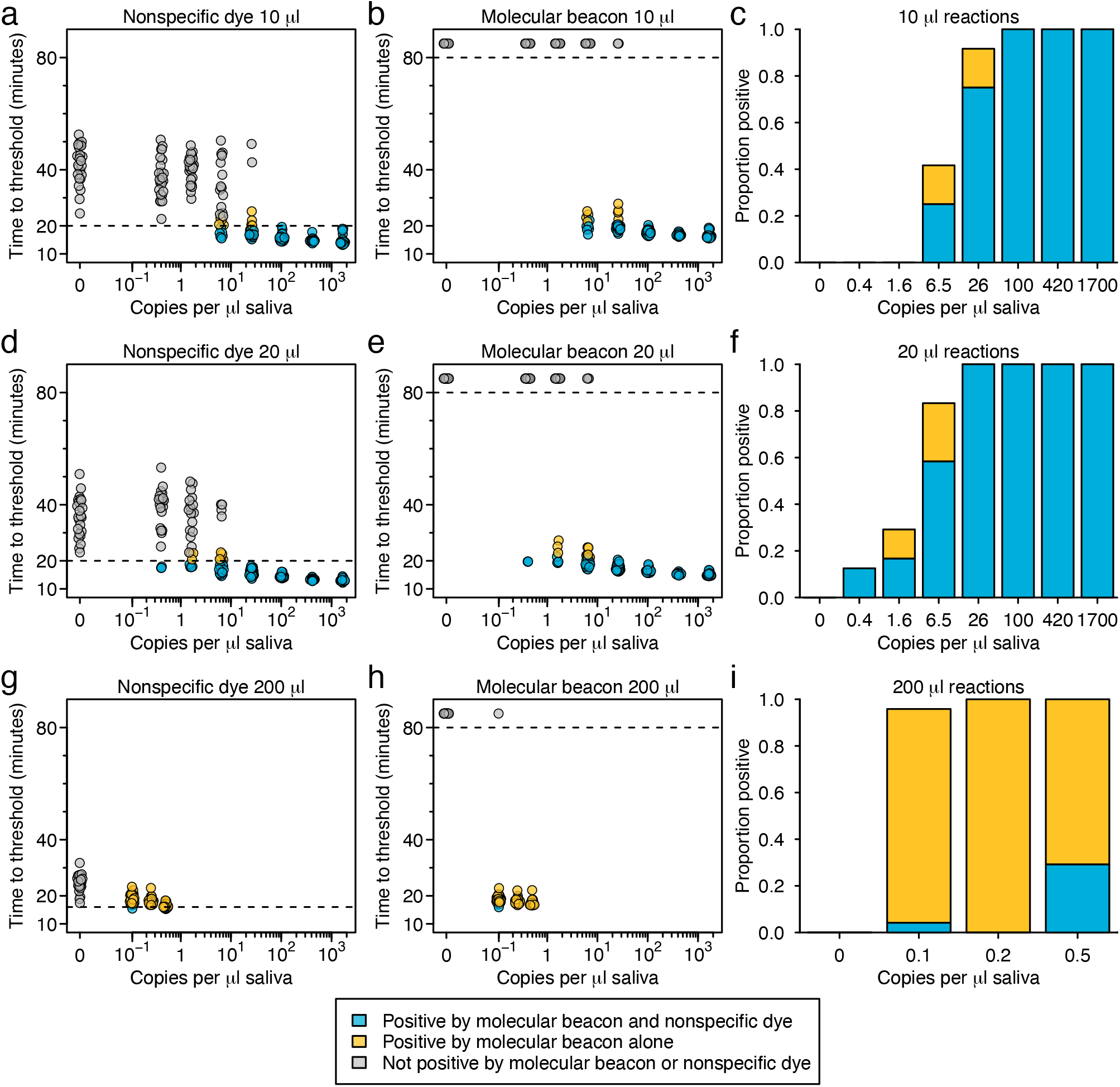
Comparison of nonspecific dye and LAMP-BEAC performance in detecting SARS-CoV-2 RNA with increased reaction volume. Times to threshold was estimated in reactions amplifying synthetic SARS-CoV-2 RNA diluted in inactivated saliva with the N2 primer set with modified LB primer in total reaction volumes of 10 μl (A-C), 20 μl (D-F) and 200 μl (G-I). Time to threshold was calculated for both nonspecific dye (A,D,G) and molecular beacon N2_LBMB_S3 labeled with a cyanine-3 fluorophore (B,E,H) within each reaction. Dashed horizontal line indicates threshold used to call a well positive or negative; less than 20 minutes for nonspecific dye in 10 or 20 μl reactions, less than 17 minutes for nonspecific dye in 200 μl reactions and 80 minutes (the time the reactions were terminated) for the molecular beacon. For ease of comparison, dots are colored blue if called positive by nonspecific dye and molecular beacon, yellow if called positive by molecular beacon alone, and grey if called negative by both methods (no sample was called positive by nonspecific dye and negative by molecular beacon). Times to threshold for molecular beacon wells with no observed increase in fluorescence were arbitrarily set to 85 minutes. Points are offset slightly on the x-axis for visualization. G-I) A comparison of the proportion of wells called positive in the reaction shown in A-B, D-E and G-H by nonspecific dye and molecular beacon (blue) or molecular beacon alone (yellow) in 10 μl (G), 20 μl (H) or 200 μl (I) reactions (n=24 for each dilution).

We thus ran a series of 200 μl reactions with 80 μl of saliva input (Fig. 5g-i). Amplification detection by non-specific dye performed poorly in these large reaction volumes, with rapid amplification observed in all negative wells. With this strong background amplification, setting a threshold to call as positive was problematic. The fastest time to threshold in a negative well was 17 minutes while the fastest time to threshold in a positive well was 15 minutes, leaving little room for discrimination (Fig. 5g). Even using the unrealistically tight threshold of calling any amplification detected in less than 17 minutes positive, nonspecific dye did not achieve high sensitivities in reactions with low copy numbers (Fig. 5i).

In contrast, the N2_LBMB_S3 molecular beacon showed almost perfect discrimination in the 200 μl reactions (Fig. 5h). No amplification was detected by molecular beacon in any of the 24 negative reactions (4800 μl of total reaction volume) over the 80 minute reaction time. In wells containing synthetic SARS-CoV-2 RNA, the molecular beacon detected 100% of reactions containing 0.5 or 0.25 copies of RNA per μl of saliva input and 23/24 reactions containing 0.1 copy of RNA per μl of saliva (Fig. 5i). Note that even with these low target concentrations, the absence of signal in negative wells means that real time quantification is not necessary and simple endpoint read out is just as discriminative.

## Discussion

Standard RT-LAMP is an attractive method for assay of SARS-CoV-2 RNA in patient samples due to the simplicity of the method and the use of a supply chain orthogonal to the clinical assay supply chain. However, conventional LAMP typically detects only the presence of amplified bulk DNA, and thus assays can be complicated by nonspecific amplification. Improved specificity can be achieved by sequence-specific detection, and multiple methods have been proposed[10-15]. Here we introduce a particularly convenient and effective method for sequence-specific detection of SARS-CoV-2 RNA in unpurified saliva using molecular beacons—LAMP-BEAC—that does not require manipulation of reaction products, can be carried out in a multiplex format in a “single tube”, greatly reduces the potential for false positives, and allows increased sensitivity.

In simple small volume assays, the LAMP-BEAC method on straight saliva may not be as sensitive as RT-qPCR on purified RNA, but it can be implemented inexpensively, potentially allowing frequent population screening. The reaction set up and incubation can be done in less than an hour with a simple heat block, allowing rapid and high throughput turnaround. The assay can even be read out visually with quite inexpensive equipment (Fig. 1c), e.g. a ∼$25 p51 viewer (from miniPCR). Thus the LAMP-BEAC assay meets the needs articulated by modeling studies for effective surveys of asymptomatic populations[1].

When greater sensitivity is needed, a simple increase in reaction volume allows LAMP-BEAC to detect SARS-CoV-2 RNA down to a 0.1 copy per μl of saliva (Fig. 5). This high sensitivity did not require purification or concentration of RNA.

Comparison of LAMP-BEAC to RT-qPCR showed better concordance between the RT-qPCR assay carried out on the same saliva samples than for RT-qPCR carried out on eluates from NP swabs. For the assays on saliva, all samples with detectable STATH or SARS-CoV-2 amplification agreed between LAMP-BEAC and RT-qPCR, suggesting that both are similarly effective at identifying samples with higher viral RNA copy numbers. The reason for divergence with some results for RT-qPCR on NP swabs is unknown—however, differences in viral RNA loads within patients at different body sites is well documented, possibly accounting for some differences[24].

Recently Vogels et al. reported SalivaDirect, an RT-qPCR assay run on inactivated but unpurified saliva [25]. SalivaDirect uses a duplex single-tube analytical method, with one amplicon targeting SARS-CoV-2 RNA and another targeting a human RNA. This parallels our triplex LAMP-BEAC targeting viral RNA and human STATH RNA (Fig. 4). The LAMP-BEAC provides an effective complement to SalivaDirect since it does not rely on commercial enzyme mixes, potentially providing more resilience to possible supply chain disruptions, and can be carried out as an end-point assay using a heat block and fluorescent viewer, thus bypassing the need for quantitative real-time PCR machines. Additionally, LAMP-BEAC allows identification of variants of concern using targeted molecular beacons [26].

A protocol based on LAMP-BEAC has received Clinical Laboratory Improvement Amendments (CLIA) approval as a laboratory developed test and has been used to screen thousands of samples per week. To date more than 40 asymptomatic subjects have been identified as positive and referred for follow up care. LAMP-BEAC thus enables rapid, affordable and scalable screening programs.

## Conclusions

Affordable, fast and robust testing is a necessity for the control of SARS-CoV-2 and future pandemics. LAMP-BEAC meets all these criteria while allowing sensitive detection when needed, all while using simple isothermal amplification and supply chains independent of commercial qPCR assays.

## Methods

### Design of Molecular Beacons

Beacons were designed to detect amplification product generated using previously published LAMP primer sets. To design beacons targeting the loop region of the LAMP product, we mapped the FIP and BIP primers to the SARS-CoV-2 genome to find the entire forward and backward loop regions of the amplicon (potentially including regions outside the original LF and LB primers). We then selected the most GC-rich subsequences within these loops and selected bases for LNA modification based on the predicted change in melting temperature using a stepwise greedy heuristic of consecutively adding the LNA with the highest predicted Tm. Additional nucleotides were then added to the 5’ and 3’ ends, avoiding strings of 4 guanine or a guanine next to the fluorophore, to form a hairpin with predicted melting temperature between 57-65 °C. Melting temperatures were predicted using OligoAnalzyer v3.1 (IDT). Where possible terminal bases of the target sequence were used as part of the hairpin. To allow easy and relatively affordable synthesis, beacons were kept shorter than 25 nt with 6 locked nucleic acids. With these constraints, commercial synthesis (IDT) provided an average yield ∼50 nmoles at a cost of ∼$400 giving a final cost per 15 μl reaction of US $0.03. Local synthesis was also implemented on a BioAutomation MerMade 4 oligonucleotide synthesizer in case of supply chain failure. Successful design often required several iterations. These earlier iterations are listed in Supplementary Table 1 and their fluorescence characteristics are compared in Supplementary Figure S3.

As an additional optimization, we created a new N2 loop primer to reduce overlap with molecular beacons. The loop primers of the N2 primer set span almost the entirety of the loop regions and thus present the possibility for false amplification involving the primer to generate targets for beacons landing in the same region. As a work around, we shortened the backward loop primer while maintaining binding affinity using locked nucleic acids (N2_LB_2, Table S1) and used this modified primer set for further testing.

### Design and purification of polymerases

We initially chose the D720A mutant of *Geobacillus stearothermophilus* PolA for LAMP due to the high strand displacement activity observed for a closely related polymerase[27]. Using strain DSM 13240, the coding sequence corresponding to the large polymerase fragment (Bst-LF; residues 290-878) was amplified from genomic DNA, ligated into CDFDuet (Novagen) in-frame with an N-terminal hexahistidine tag and the D720A substitution was incorporated. To explore alternative Bst-LF variants for LAMP, we generated the R433A and R433P variants, each of which results in disruption of the salt-bridge formed with Asp720 in the wild-type enzyme (protein data bank: 1XWL).

Bst-LF was expressed in strain BL21(DE3) at 37 °C with 2xYT medium and IPTG induction for 3 h. Pelleted cells were stored at -70 °C prior to purification. Cells were lysed using an Avestin cell disrupter in 50 mM sodium phosphate, pH 8, 300 mM NaCl, 2 mM MgCl2, 5 mM 2-mercaptoethanol, and protease inhibitors. After centrifugation, the cleared lysate was purified at 4 °C using a 5 ml Talon column (Clontech) following the manufacturer’s protocol. Eluted fractions from Talon were diluted 1:1 with buffer HepA (20 mM Tris-HCl, pH 7.4, 5 mM MgCl_2_, 10 mM 2-mercaptoethanol) and purified on an 8 mL heparin sepharose column (GE), and at 20 °C using a 0-500 mM NaCl gradient. Heparin fractions were pooled and dialyzed overnight vs 20 mM Tris-HCl, pH 8, 50 mM NaCl, 10 mM 2-mercaptoethanol followed by anion exchange chromatography at 20°C using an 8 ml MonoQ (GE) column. Bst-LF eluted as sharp peaks from a 15-30% sodium chloride gradient in buffer containing 20 mM TrisHCl, pH 8, 10 mM 2-mercaptoethanol. Purified Bst-LF mutants were concentrated using Millipore centrifugal concentrators, glycerol added to 10%, and aliquots were flash frozen and stored at -80 °C. Primer extension assays using M13 DNA template and ^3^H-dTTP labeled dNTPs were used to establish specific activity as described for commercially prepared Bst (NEB).

To demonstrate that RT-LAMP can be performed using a reverse transcriptase generated in-house, we first constructed a synthetic gene for the HIV1 RT p66 (strain NL4-3) subunit containing substitutions expected to confer thermal stability (RTx; NEB). The p66 sequence was inserted into pET29b and the p51 subunit coding sequence was amplified by PCR and inserted in frame with an N-terminal hexahistidine tag in CDFDuet. An alternative RT (RT2m) was produced using a similar approach with HIV2 RT as the template (Genbank AAB25033), where thirteen naturally occurring substitutions were incorporated. The full-length subunit was inserted into pCDFDuet and the smaller subunit was fused to a C-terminal hexahistidine tag after Thr436 in pETDuet.

For both RTs, the subunits were co-expressed in BL21(DE3) cells and 2xYT media, with IPTG induction at 20 °C for 5h. Pelleted cells were stored at - 70 °C prior to purification. Cells were lysed using an Avestin cell disrupter in 50 mM sodium phosphate, pH 8, 300 mM NaCl, 2 mM MgCl_2_, 5 mM 2-mercaptoethanol, and protease inhibitors. After centrifugation, the cleared lysate was purified at 4 °C using a 5 ml Talon column following the manufacturer’s protocol. Eluted fractions from Talon were dialyzed vs buffer A (20 mM Tris-HCl, pH 7.4, 50 mM NaCl, 5 mM MgCl_2_, 10 mM 2-mercaptoethanol) and purified on an 8 mL heparin sepharose column at 20 °C using a 150-400 mM NaCl gradient in buffer A. Heparin fractions were pooled and concentrated on 5 ml Ni-NTA resin (Qiagen), followed by elution with 20 mM TrisHCl, 100 mM NaCl, 10 mM 2-mercaptoethanol, 250 mM imidazole, pH 8. Purified RT was dialyzed vs 20 mM Tris-HCl pH 8, 50 mM NaCl, 10 mM 2-mercaptoethanol and concentrated using Millipore centrifugal concentrators. Glycerol was added to 10%, and aliquots were flash frozen and stored at -80 °C. Primer extension assays using poly-A template and ^3^H-labeled dTTP were used to determine specific activity at 50 °C as described for commercial RTx (NEB).

### RT-LAMP reaction mixtures

RT-LAMP reactions were prepared by mixing 7.5 μl commercial 2x LAMP master mix (NEB E1700L) or our own LAMP mix (40 mM TrisHCl, pH 8.5, 20 mM (NH4)_2_SO4, 100 mM KCl, 16 mM MgSO4, 0.2% Tween-20, 2.8 mM each dNTP, 16 µg/ml polA LF, and 2.6-7.7 µg/ml RT) with 1.5 μl of 10x primer/beacon master mix (final concentration: 1.6 μM FIP/BIP, 0.2 μM F3/B3, 0.4 μM LF/LB, 0.25 μM beacon) and 6 μl of sample and/or water. Larger 20 and 200 μl and smaller 10 μl reactions were scaled proportionally. For multiplexed LAMP reactions, the final total concentration of primers was preserved while maintaining the individual beacon concentrations e.g. when two primer sets were added to the same reaction, the individual primer concentrations were halved while beacon concentrations remained at 0.25 μM. For non-specific amplification detection, LAMP fluorescent dye (NEB B1700S) was used at a 100-300x final dilution.

### Assays using LAMP-BEAC

LAMP-BEAC reactions were performed at 60-65 °C with fluorescent quantification every 30 seconds on a ThermoFisher QuantStudio 5 or ThermoFisher QuantStudio 6. Reactions typically completed within 45 minutes but for research purposes data was collected for additional time spans. The synthetic SARS-CoV-2 RNA used as a standard during assay development was obtained from Twist (MT007544.1). After reaction completion, for melt curve analysis, the reaction was heated to 95 °C for 5 minutes to inactivate any remaining enzyme, cooled to 25 °C (at a rate of 0.2 °C/sec) and then slowly heated to 95 °C with fluorescence measured every degree.

Time to threshold was calculated as the time required for fluorescence from nonspecific dye detected at 520 nm to reach 200,000 relative fluorescence units (RFU) for 10 or 20 μl reactions higher than baseline or 400,000 RFU for 200 μl reactions. For fluorescence from molecular beacon detected at 587 nm, time to threshold was calculated as the time required to reach 20,000 RFU higher than baseline for 10 or 20 μl reactions or 40,000 RFU for 200 μl reactions. Baseline was calculated as the average RFU over the first 2.5 minutes of the reaction for 10 or 20 μl reactions or the average over the second 2.5 minutes for 200 μl reactions to allow the reactions to completely equilibrate.

### RT-qPCR to characterize saliva samples

RNA was extracted from ∼140 μl saliva using the Qiagen QIAamp Viral RNA Mini Kit. The RT-qPCR assay used the CDC 2019-nCoV_N1 primer-probe set (2019-nCoV_N1-F: GACCCCAAAATCAGCGAAAT, 2019-nCoV_N1-R: TCTGGTTACTGCCAGTTGAATCTG, 2019_nCoV_N1-P: FAM-ACCCCGCATTACGTTTGGTGGACC-IBFQ). The RT-qPCR master mix contained: 8.5 μl dH_2_O, 0.5 μl N1-F (20 μM), 0.5 μl N1-R (20 μM), 0.5 μl N1-P (5 μM), 5.0 μl TaqMan™ Fast Virus 1-Step Master Mix per reaction. 5 μl of extracted RNA was added to 15 μl of prepared master mix for a final volume of 20 μl per reaction. Final concentrations of both 2019-nCoV_N1-F and 2019-nCoV_N1-R primers were 500nM and the final concentration of the 2019-nCoV_N1-P probe was 125nM. The assay was performed using a ThermoFisher QuantStudio 5. The thermocycler conditions were: 5 minutes at 50 °C, 20 seconds at 95 °C, and 40 cycles of 3 seconds at 95 °C and 30 seconds at 60°C.

## Data Availability

The data supporting the findings of this study are available within the article.

## Abbreviations

RT-qPCR: reverse transcription quantitative polymerase chain reaction
COVID-19: coronavirus disease from 2019
SARS-CoV-2: severe acute respiratory syndrome coronavirus number two
RT-LAMP: reverse transcription loop-mediated isothermal amplification
RFU: relative fluorescence units

## Declarations

### Ethics approval and consent to participate

All sample collection was carried out under IRB-approved protocols (IRB protocol #842613 and #813913). Salivary samples were collected from possible SARS-CoV-2 positive patients at one of three locations: (1) Penn Presbyterian Medical Center Emergency Department, (2) Hospital of the University of Pennsylvania Emergency Department, and (3) Penn Medicine COVID-19 ambulatory testing center. Inclusion criteria including any adult (age>17 years) who underwent SARS-CoV-2 testing via standard nasopharyngeal swab at the same visit. Patients with known COVID-19 disease who tested positive previously were excluded. After verbal consent was obtained by a trained research coordinator, patients were instructed to self-collect saliva into a sterile specimen container which was then placed on ice until further processing for analysis.

### Consent for publication

All authors have reviewed the manuscript and consented to allow publication.

### Availability of data and materials

All data newly generated in this study is disclosed in the published manuscript.

### Authors contributions

SS-M, BDA, RGC, GVD and FDB designed the study; JG-W, LJT, BSA, and RGC collected clinical specimens; SS-M, YH, AMR, AGl, AGa, LH, PD, CN, AK and GVD carried out assays; SRW and YL grew SARS-CoV-2 in tissue culture; SS-M, GVD, and FDB wrote the paper.

## Acknowledgements

We are grateful to members of the Bushman, Van Duyne and Collman laboratories for help and suggestions. We acknowledge the assistance of the Penn Medicine BioBank, including JoEllen Weaver and Daniel Rader. This work was supported by the Penn Center for Research on Coronaviruses and Other Emerging Pathogens.

## Supplementary Material

Table S1. Oligonucleotides used in the LAMP-BEAC assay.

Table S2. Clinical samples and results of assays for SARS-CoV-2 and human RNA.

Fig. S1. Reaction progression and melt curves for LAMP-BEAC reactions carried out on clinical saliva samples. Each column represents a single sample assayed. Sample names are as in Table S2. Heavy boxes indicate samples called positive for that amplicon.

Fig. S2. Comparison of laboratory-purified and commercial polymerase enzymes using a quadruplex LAMP-BEAC assay of saliva samples spiked with synthetic SARS-CoV-2 RNA. Assays were carried out using an amplicon to detect human STATH RNA (A) and three amplicons to detect SARS-CoV-2 (B-D). For these assays, synthetic SARS-CoV-2 RNA was diluted into saliva (inactivated as described[9]); copies per microliter are shown by the color code in the lower right. For A-D, the x-axis shows time after starting the assay, and the y-axis shows fluorescence intensity. E-H shows melt curve analysis for samples in A-D. For E-H, the x-axis shows temperature, and the y-axis shows fluorescence intensity. (I-P) Assays are exactly as in A-H, but commercial reverse transcriptase and DNA polymerase (NEB Warm Start LAMP Kit master mix product number E1700L) were used instead of the locally designed and purified polymerases.

Fig. S3. Testing of the various iterations of molecular beacons reported in Table S1. A) The amplification and melt curves of various SARS-CoV-2 targeted beacons. Each plot shows amplification and melt curves in reactions amplifying saliva (grey) or saliva doped with 10,000 copies of Twist synthetic SARS-CoV-2 RNA (red). Beacon and fluorophore identity is indicated above each plot. B) As in A but for beacons targeting human sequences. Amplification was performed on saliva from two different individuals (yellow or orange) or water (black).

