## Supplementary figures and images for "LAMP-BEAC: Detection of SARS-CoV-2 RNA Using RT-LAMP and Molecular Beacons"

### Supplemental Figure 1

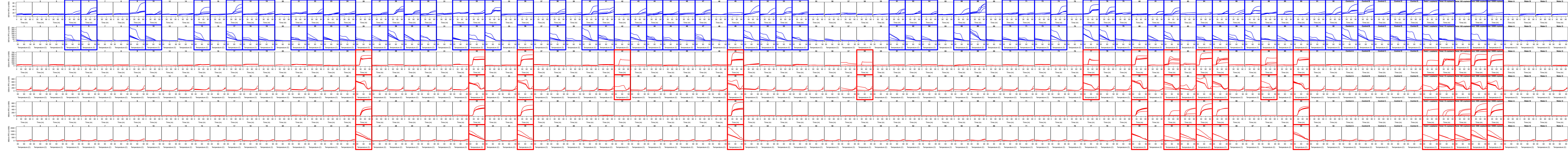

### Supplemental Figure 2

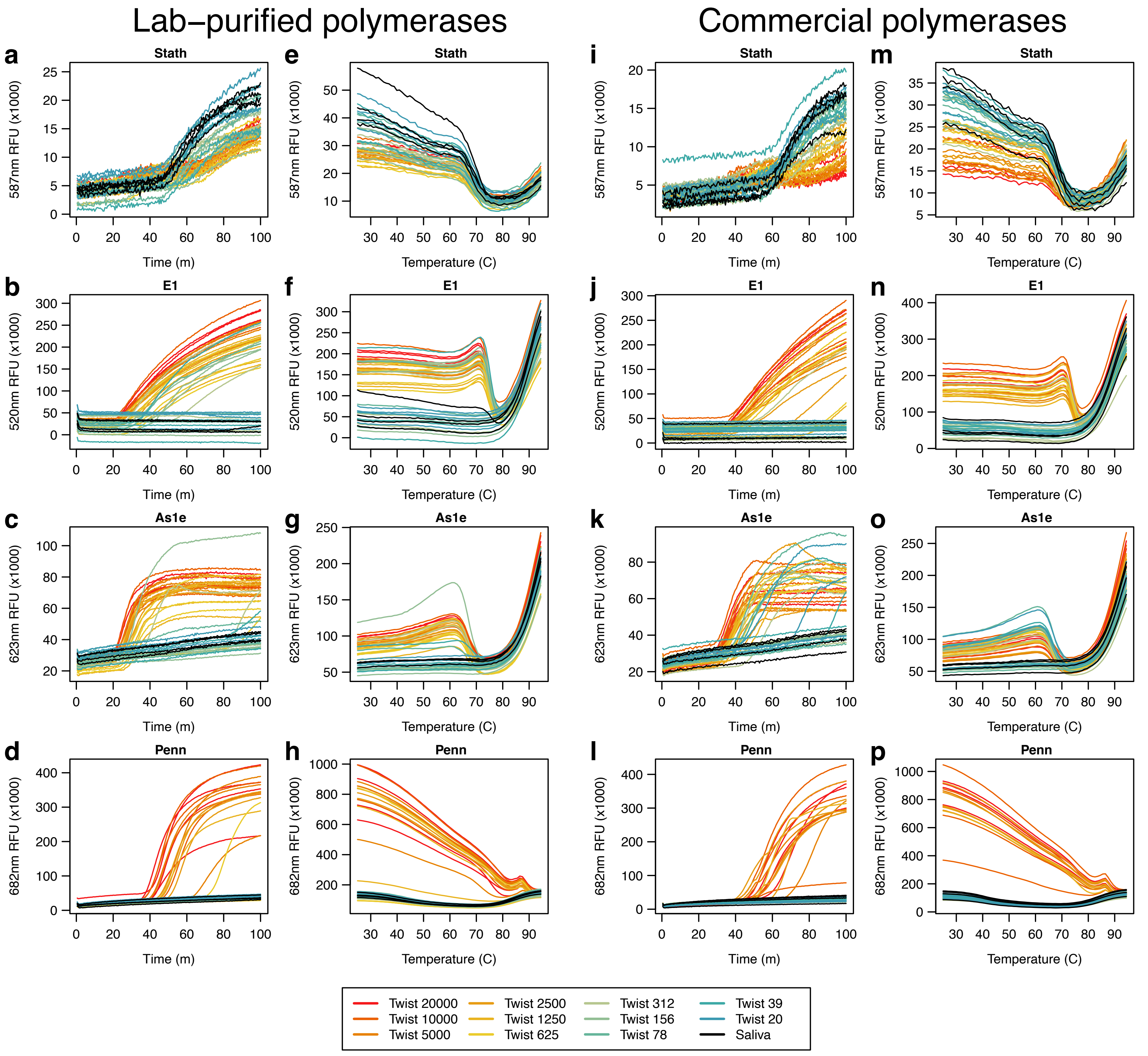

### Supplemental Figure 3

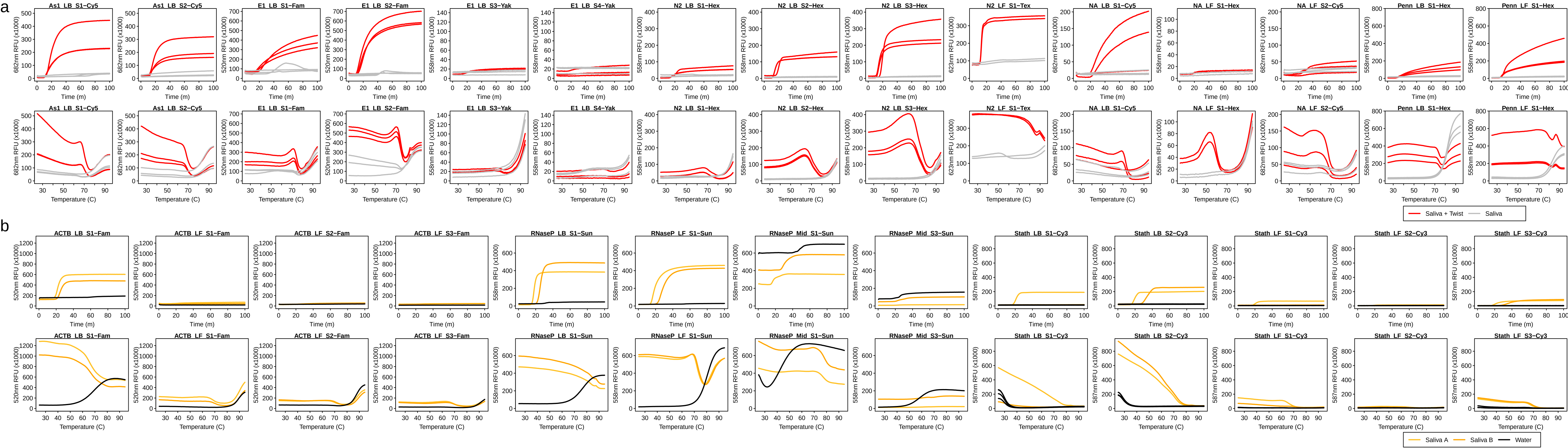
